# Biomarker Concordance Between Dried Plasma Spots of Capillary Blood and Venous Plasma in Traumatic Brain Injury: An Analytical Feasibility Pilot Study

**DOI:** 10.64898/2026.09.16.26361861

**Authors:** Devin Jackson, Katie Tehas, Kristy Radeker, Anthony DeLizza, Caroline Popper, Elaine Peskind, Ava Puccio, Raquel C. Gardner, John B. Williamson, Abigail B. Waters, Lisa H. Merck, Geoff Manley, Kevin K. Wang, William E. Haskins

## Abstract

**Background:** Traumatic brain injury (TBI) increases long-term risk for cognitive decline and neurodegeneration, motivating objective blood biomarkers that can be measured repeatedly over time. However, reliance on venipuncture limits longitudinal monitoring, particularly outside clinic settings.

**Objective:** We conducted a pilot feasibility study comparing fingerstick capillary blood collected as dried plasma spots (DPS) with matched venous EDTA plasma for quantification of glial fibrillary acidic protein (GFAP) and neurofilament light (NfL).

**Methods:** Paired samples were collected prospectively across matched time points spanning hours to decades post-injury and assayed using high-sensitivity immunoassays.

**Results:** Absolute protein recovery from DPS was lower and more variable than venous plasma, but paired GFAP and NfL measurements remained well-correlated. We further evaluated the GFAP/NfL ratio as an internal, sample-based normalization strategy and observed improved concordance between capillary DPS and venous plasma compared with either analyte alone.

**Conclusions:** These pilot findings support the analytical feasibility of capillary DPS sampling for GFAP and NfL measurement in TBI and identify the GFAP/NfL ratio as an exploratory internal-normalization approach. Formal analytical and clinical validation — including outcome linkage and prospective evaluation — will be required before capillary DPS can be considered for clinical monitoring applications.

**Highlights:**

- First high-frequency (twice-daily for 14 days) paired capillary-DPS / venous-plasma GFAP and NfL trajectories in acute TBI (Cohort #1, n = 4).
- Strong within-subject capillary-venous correlations for GFAP (ρ up to 0.647) and NfL (ρ up to 0.902); the GFAP / NfL ratio reaches ρ = 0.797–0.977.
- Cross-sectional capillary-DPS biomarkers separate acute, subacute, and chronic TBI populations across a multi-site cohort (Cohort #2).
- Concurrent work by Bouthors et al. (CCLM 2026) supports prioritizing GFAP and NfL over UCH-L1 for capillary panels because UCH-L1 is intra-erythrocytic and hemolysis-sensitive.
- Capillary DPS sampling establishes analytical feasibility for paired capillary–venous GFAP and NfL measurement; formal validation of context-of-use, unsupervised collection, and outcome linkage is required before clinical deployment.

## Introduction

### Traumatic Brain Injury (TBI)

TBI is an urgent unmet medical need. In 2021, there were approximately 21 million incident cases and 38 million prevalent cases of TBI globally.^1^ TBI is also a signature injury among US military personnel in modern warfare, with more than 514,583 confirmed cases from 2000–2024.2 More than 185,000 veterans who use the U.S. Department of Veteran Affairs for their health care have been diagnosed with at least one TBI.3 Recent statistics from the U.S. Centers for Disease Control and Prevention indicate that there are ∼ 214,000 TBI-related hospitalizations in the U.S. annually—representing 8% of all of the 2.8M new TBIs and 68% of the $80B hospitalization costs in the U.S. annually.^4^ Survivors of TBI, particularly moderate-severe TBI (msTBI) patients defined by the Glasgow Coma Scale (GCS) score of 3-12,5 can suffer long-term or life-long health problems. Fall, firearm-related injuries, and motor vehicle accidents lead to most msTBIs, and msTBI patients are the most likely to be readmitted within 30 days of hospital discharge. This is particularly true for older patients due to falls and co-morbidities.^6–16^ For example, 8.9% of 135,342 TBI patients hospitalized in 2014 were readmitted within 30 days of discharge: age, documentation of a fall, and intentional self-injury at the index admission were positively associated with readmission.^16^ Lastly, in addition to the impact to the individual and their families, TBI incurs large societal and economic costs.^17^

### The Post-Acute Monitoring Barrier

In 2018, based on the ALERT-TBI study,18 the FDA authorized the first acute mild TBI (mTBI or concussion) blood test for neuronal ubiquitin C-terminal hydrolase-L1 (UCH-L1) and astrocytic glial fibrillary acidic protein (GFAP) — the Banyan Brain Trauma Indicator — through the De Novo pathway to reduce the need for computed tomography (CT) brain scan-based diagnosis within 12 hours of injury.^19–21^ A subsequent point-of-care implementation of the GFAP + UCH-L1 plasma panel on the Abbott i-STAT Alinity TBI cartridge received 510(k) clearance in 2021.51-53 This test fills an important gap in diagnosis of mTBI by aiding in the identification of brain lesions and reducing the need for CT scans. Neurofilament light chain (NfL) has emerged as another promising blood-based biomarker of TBI, reflecting axonal damage and demonstrating strong diagnostic and prognostic utility across injury severities51,53-55. Unlike GFAP and UCH-L1 which have peak levels within 24- 48 hr post–injury, NfL show a delayed rise in TBI patients and peaking at about 3 weeks post injury54. However, to date, there are no FDA-cleared diagnostic medical devices to monitor post-acute blood-based biomarkers of msTBI or mTBI beyond 12h, nor any blood tests that can trace the unique recovery trajectory of each TBI patient from the acute to post-acute phases. This is a significant barrier to TBI patients obtaining meaningful follow-up care and management in hospital and post-hospital rehabilitative settings.

The clinical importance of blood-based TBI biomarkers has recently been formalized at the framework level. The NIH-NINDS TBI Classification and Nomenclature Initiative proposed replacing the severity-only Glasgow Coma Scale paradigm with a multidimensional characterisation built on four pillars — Clinical, Biomarker, Imaging, and Modifiers (CBI-M) — in which blood-based measures such as GFAP and NfL constitute a core diagnostic pillar.47 Situating our work within CBI-M, the present study addresses the practical bottleneck of the biomarker pillar: how these measures can be sampled repeatedly, minimally invasively, and outside the clinic. By establishing analytical feasibility for capillary dried-plasma-spot GFAP and NfL, we aim to extend the reach of the CBI-M biomarker pillar from the acute, venous, in-hospital setting toward longitudinal and remote post-acute monitoring.

### The Capillary Blood Sampling Gap

Blood-based biomarkers are more advantageous than CSF biomarkers because blood can be sampled less invasively than lumbar puncture needed for sampling CSF and because recent advances in immunoassay sensitivity and specificity have largely overcome the analytical challenge of quantifying the lower levels of CNS biomarkers (and higher levels of more abundant proteins) in blood than CSF.

Venipuncture blood sampling. State-of-the-art assays are available for GFAP, UCH-L1 and other biofluid biomarkers with venipuncture blood sampling to collect plasma and serum.18,20,21 However, although venipuncture sampling is less invasive than CSF sampling, it is still burdensome to patients and caregivers. The need for venipuncture, as well as cold-chain transport of blood samples, hampers widespread adoption of standard biomarker platforms, limiting use in post-acute brain health monitoring. This is particularly true for elderly patients, patients who lack mobility or live in more remote areas where visits to clinics (neurology, memory, or dementia clinics) are more problematic, and for high-risk patients who may be unable or unwilling to present to a hospital in person for logistical or medical/social reasons. Remote and dried-matrix sampling is also enabling for high-frequency monitoring in austere military environments and rural or remote civilian locations, for sample banking supporting add-on biological investigations in interventional trials, for pandemic-era public-health scenarios such as those seen during the COVID-19 pandemic, and following mass casualty events. However, it is also true in general for large well-funded and highly informative clinical research studies.26,44

#### Capillary blood sampling

Unlike conventional venipuncture blood sampling, fingerprick sampling of capillary blood is minimally invasive, making it ideal for routine (serial) blood sampling and follow-up studies. To date, there are no FDA-cleared capillary blood tests for biomarkers of TBI or adjacent brain conditions. This is surprising, given the ubiquity of dried (whole) blood spots for capillary blood sampling as part of prenatal testing, reports of longitudinal studies for non-CNS indications,^27, 28^ and other situations where minimally invasive sampling is ideal. However, there is sometimes a poor correlation between protein biomarker levels in dried blood spots (D<u>B</u>S) due to irreversible adsorption and poor or irreproducible recovery.^29^ Recently, several device manufacturers have met this challenge by sampling 2-4 drops (∼40-80 µL) of fingerprick capillary blood to reproducibly collect ∼3-10 µL of plasma onto a lightweight, inexpensive, paper-based dried plasma spot (D<u>P</u>S) sampling device for ambient temperature transport of samples to the laboratory for analysis. Some of these devices are similar to a successful DBS sampling device that is capable of measuring analytes with volumetric precision from varying applied sample volumes and hematocrit levels.^30,37^

### This Study

To address the post-acute monitoring barrier and reduce the biosampling burden, we evaluated the analytical feasibility and preliminary clinical relevance of capillary blood sampling for TBI biomarkers. We prospectively collected paired DPS capillary blood and venous EDTA plasma from TBI patients at matched time points. Samples underwent cold-chain transport for high-sensitivity immunoassay quantification of GFAP and NfL. Here, we present the trajectory profiles and correlations of GFAP and NfL between these paired sampling methods, demonstrating the feasibility of capillary blood as a minimally invasive sampling approach for paired biomarker measurement for TBI and related neurodegenerative conditions.

## Methods

### Study Design and Patient Cohorts

This study evaluated two prospective clinical cohorts (**Table 1 and 2**).

**Table 1.** Demographic and clinical characteristics of the four civilian acute TBI subjects (Prospective Cohort #1, n = 4) enrolled for high-resolution paired capillary dried plasma spot (DPS) and venous EDTA plasma sampling (up to twice daily for 14 days). Reported per subject: sex, age range, Glasgow Coma Scale (GCS), and TBI severity classification (mTBI: GCS 13–15; msTBI: GCS ≤ 12).

| Subject ID | Sex | Age range | GCS |
| --- | --- | --- | --- |
| Subject A | M | 66–70 | 13 |
| Subject B | M | 41–45 | 14 |
| Subject C | M | 21–25 | 7 |
| Subject D | M | 61–65 | 8 |

**Table 2.** Demographic and clinical characteristics of the cross-sectional TBI cohort (Prospective Cohort #2, n = 29) enrolled across three sites and three injury phases: acute geriatric civilian mTBI (UCSF), subacute / chronic geriatric civilian TBI (Puget Sound VA), and chronic military / Veteran TBI (UF–VA). Reported per sub-cohort: sex distribution, mean age and GCS (± SD), and key clinical features.

| Subject Type | Sex | Age (± SD) | GCS (± SD) | Clinical Characteristics |
| --- | --- | --- | --- | --- |
| **Geriatric Civilian Acute mTBI (n = 14)** | 7 M, 7 F | 76.1 ± 5.7 | 14.7 ± 0.5 | 28.5% CT + |
| **Geriatric Civilian and Veteran Subacute/Chronic mTBI (n = 10)** | 7 M, 3 F | 73.7 ± 5.7 | 8-12 (n=5), 13-15 (n=5) | 40% CT + 70% falls, 20% motorized vehicle accident |
| **Military Chronic mTBI (n = 5)** | 4 M, 1 F | 47.8 ± 3.3 | 13-15 | 2 CDR = 0; 2 CDR = 0.5; 1 CDR undefined |
| **Controls (n = 21)** | 10 M, 11 F | 73.3 ± 5.2 | N/A | N/A |

For Prospective Cohort #1, Institutional Review Board (IRB) approval was obtained at Virginia Commonwealth University (VCU) under Advarra protocol Pro00070062 to collect paired dried plasma spot (DPS) fingerstick samples and venous EDTA plasma. Samples were collected up to twice daily for 14 days post-injury (or until hospital discharge) from patients with moderate-to-severe TBI (msTBI; Glasgow Coma Scale [GCS] 3–12) and mild TBI (mTBI; GCS 13–15). Representative analyses included 113 paired samples from two mTBI subjects (Subjects A and B) and 100 paired samples from two msTBI subjects (Subjects C and D), alongside single-timepoint control samples. Patient demographics are summarized in **Table 1**.

For Prospective Cohort #2, multi-site IRB approval facilitated the collection of paired, matched-timepoint samples across diverse patient profiles (Table 2) under the following site approvals: University of California San Francisco (UCSF) protocol 19-28925, which also served as the IRB of record for the University of Pittsburgh under a reliance agreement ceding review (UPitt local protocol STUDY20110253); University of Florida protocol IRB202102196 (North Florida Foundation for Research and Education; University of Florida / Malcolm Randall Veterans Affairs Medical Center site); and VA Puget Sound IRB 2 protocol 1653649-1 (Seattle Institute for Biomedical and Clinical Research; VA Puget Sound site). These included acute geriatric civilian mTBI subjects (University of California San Francisco and University of Pittsburgh), n = 14; subacute/chronic geriatric civilian and Veteran mTBI subjects (University of Florida/Malcolm Randall Veterans Affairs), n = 10; and Veteran chronic mTBI and non-TBI subjects (Veterans Affairs Puget Sound), n = 5 [Cohort #2 total n = 29]. Representative data comprised 31 paired DPS and venous plasma samples from 24 geriatric civilian and Veteran subjects (acute: <24 hours post-injury, n = 8; subacute: 1 week to 3 months, n = 11; chronic: 1 year ± 45 days, n = 3) and five additional Veteran subjects from the VA Puget Sound site.

### Clinical Sampling Protocols

To ensure optimal capillary blood flow, subjects washed their hands in warm water for 30–60 seconds, after which the skin was sterilized with an alcohol swab. Patients rested their hands below heart level, and a high-flow lancet (BD-366594) was used to puncture the fingertip. The first hanging drop of blood was discarded, and subsequent drops were loaded onto one of two paper-based DPS devices (Figure 5). Device #1 accommodated ∼50 µL of capillary blood (approximately two drops) to elute ∼3 µL of plasma onto a DPS disc. Device #2 accommodated ∼70 µL (3–4 drops) to elute ∼10 µL of plasma. All DPS discs were air-dried at ambient temperature for one hour before being stored individually in sealed plastic bags with desiccant at −80°C. Matched venous whole blood was concurrently drawn into K2EDTA tubes to isolate reference plasma.

### DPS Rehydration and Immunoassay Quantification

Frozen DPS discs were extracted using tweezers and placed into Corning Spin-X Centrifugal Filter Units (Fisher Scientific). Discs were rehydrated with 12 µL of a 0.1% Tween-20 detergent solution and agitated at 250 rpm for 2 hours at 4°C. Following centrifugation (3,000 rcf for 7 minutes at 4°C), the resultant eluent was diluted 1:2 in assay buffer to yield a final 1:10 dilution with a recovery volume of 12–15 µL. Biomarker levels were quantified using the Meso Scale Diagnostics (MSD) Quickplex SQ 120MM instrument with the Neurology Panel 1 kit. Samples underwent an average of 1–2 freeze-thaw cycles prior to analysis.

Statistical analyses were performed in IBM SPSS Statistics.^31^ Spearman rank correlations were used to quantify concordance between paired capillary and venous measurements, and Pearson correlations were used for select linear comparisons. Two-sided P values < 0.05 were considered statistically significant.

#### Statistical framing

The prespecified primary objective of this pilot was to characterize paired capillary–venous concordance for GFAP and NfL. Analysis of the GFAP/NfL ratio was a secondary, exploratory analysis motivated by the observed capillary–venous recovery variability. All reported p-values are descriptive; no adjustment for multiple comparisons was performed, and inferential statistics are exploratory in light of the pilot sample size and the temporal autocorrelation introduced by twice-daily longitudinal sampling in four subjects.

## Results

### Brain Injury Blood Biomarker Trajectories and Protein Recovery (high resolution twice daily sampling Cohort #1)

Brain injury blood biomarker trajectories were successfully elucidated with both capillary blood sampling in DPS device and venous plasma sampling in a set of four TBI patients (**Table 1**, **Figure 1**). Capillary DPS blood sampling and conventional venous plasma testing both successfully captured temporal brain biomarker trajectories in mild TBI/mTBI (GCS 13-15) and moderate-severe TBI/msTBI (GCS 3-12) patients. Using high-frequency matched sampling (up to twice daily for 14 days), we generated comparative profiles for two mTBI patients (Subjects A [male, age 66–70] and B [male, age 41–45]) and two msTBI patients (Subjects C [male, age 21–25] and D [male, age 61–65]) (**Figure 1A-L**).

**Figure 1.**
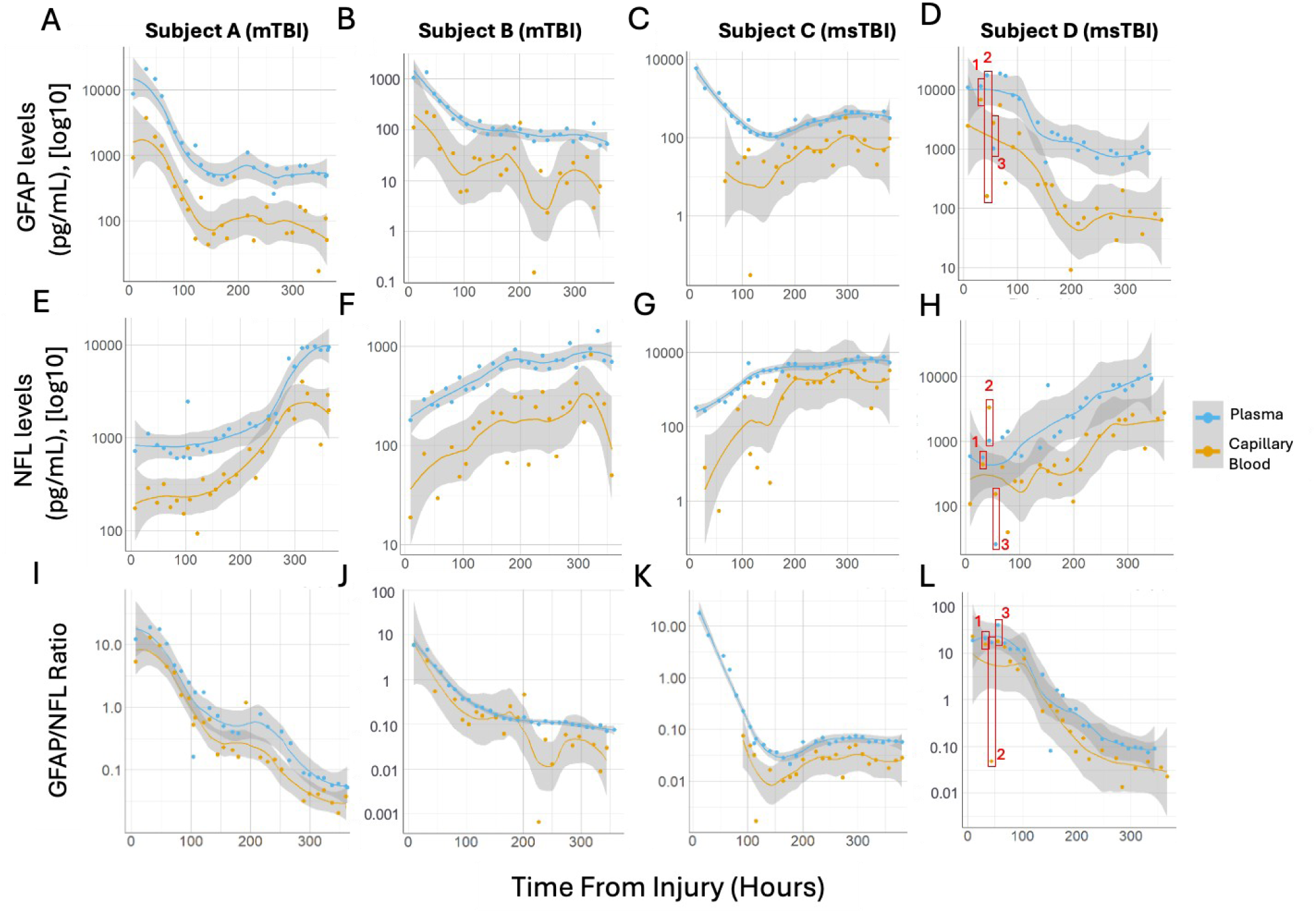
Trajectory analysis of biomarker levels vs. time for paired venous plasma and capillary blood samples from mTBI and msTBI patients from Prospective Cohort #1. Results from venous plasma and capillary blood samples are shown for GFAP and NfL levels with daily sampling for the first 48 hours and twice daily sampling thereafter until 14 days post injury (dpi). **(A, B, E, F)** Subjects A and B have mTBI (GCS = 13 and 14, respectively), and **(C, D, G, H)** Subjects C and D have msTBI subjects (GCS = 7 and 8, respectively). GFAP biomarker levels **(A-D)**, and NfL biomarker levels **(E-H)**, are displayed longitudinally. The GFAP/NfL ratio for each patient is shown for each patient **(I-L)**. **(D, H)** Timepoints boxed in red highlight clinical events: 1 – intracranial pressure (ICP) device insertion, 2 – seizure, 3 – ∼12 hours post-seizure. Longitudinal monitoring of biomarker levels in paired samples is presented in absolute values (pg/mL) versus time from injury (hours). A trendline (loess, 95% confidence interval) is overlaid for each trajectory.

Consistent with published data,^56^ GFAP biomarker levels measured in both capillary DPS and venous plasma generally followed the canonical temporal profile, with plasma levels highest within the first 24 hours of injury and tapering until approximately 6 days post-injury (dpi), where most patient curves flattened for the remainder of the sampling period (Figure 1A, B, D). The exception was Subject C, where GFAP levels slowly increased from 180 hours after the initial drop (Figure 1C). Capillary blood samples yielded overall lower GFAP levels than their plasma counterparts but followed a similar temporal trend. For plasma GFAP across all four patients, the highest levels (11,771.51 ± 4,818.70 SEM pg/mL, n=4) were observed at admission, with decay to lower levels (426.81 ± 167.31 pg/mL) by Day 14, exhibiting the most rapid decay in the first 6 dpi. It is important to note that the GFAP trajectories in capillary DPS and venous plasma samples for each subject are parallel to each other (Figure 1).

NfL levels for all four patients followed the expected increasing temporal profile for both capillary DPS and venous plasma samples (refs ^51,54,55^) (Figure 1E-H). Biomarker levels in plasma and capillary blood were lowest at the first measurement post-injury and slowly increased throughout the sampling period until 14 dpi. Capillary blood followed a similar trend as plasma, but with more variability indicated by wider 95% confidence intervals. For plasma NfL, the lowest levels generally occurred upon enrollment (453.1 ± 122.7 SEM pg/mL for plasma; 77.4 ± 39.3 SEM pg/mL for capillary blood, n=4), with the highest levels generally at 14 dpi (6,217.4 ± 2,064.9 SEM pg/mL for plasma; 2,030.2 ± 716.0 SEM pg/mL for capillary blood, n=4).

As anticipated, absolute protein biomarker recovery from the capillary DPS devices was lower when compared with venous plasma. Across the subset, mean capillary recovery was 21.7% (± 1.22% SEM, n=4) for GFAP and 40.8% (± 7.14% SEM, n=4) for NfL relative to plasma concentrations. Recovery rates remained consistent across injury severities. One exception was Subject D, in whom two early NfL capillary values exceeded matched venous plasma concentrations by more than three-fold; these values were temporally coincident with a documented seizure event, but with a single subject we treat them as pre-analytical outliers and cannot infer a causal relationship to seizure activity.

To compensate for the lower individual biomarker (GFAP and NfL) recovery observed in capillary DPS samples compared with their venous counterparts, we implemented a novel normalization approach. Specifically, we calculated the GFAP/NfL ratio for both sampling types, based on the rationale that GFAP and NfL levels within each sample (DPS and venous plasma) could serve as internal controls for one another. We indeed found that the GFAP/NfL biomarker ratio smoothed the temporal curve and reduced the difference between venous plasma and capillary samples for all patients to the point that the two corresponding curves overlap with each other (Figure 1I-L). Because GFAP is a marker of acute injury peaking in the first 24–36 hours post-injury, and NfL is a marker of subacute injury progression, a downward trending profile was observed in all subjects. Capillary blood ratios remained closer to their venous plasma counterparts compared to individual GFAP and NfL measurements. Interestingly, Subject B (Figure 1J), which had the lowest biomarker levels, also had the lowest ratios, peaking at 5.90 a.u. for venous plasma and 5.97 a.u. for capillary blood, compared to venous plasma in Subjects A, C, D (23.29 ± 8.33 a.u., n=3) and capillary blood (10.81 ± 6.56 a.u., n=3). To the best of our knowledge, this is the first study using GFAP/NfL ratio as a biomarker readout for brain injury and related neurodegenerative conditions (e.g. MCI, AD).

### Correlation of Capillary and Venous Biomarker levels (Cohort #1)

We further interrogate the correlation of capillary DPS and venous GFAP, NfL levels and GFAP/NfL ratio. Despite recovery variability, temporal trajectories of GFAP and NfL demonstrated moderate to strong correlations between capillary and venous plasma within individual subjects (Figure 2). When plotted on a logarithmic scale, paired GFAP measurements yielded Spearman rank coefficients (ρ) ranging from 0.447 to 0.647, with Subject A exhibiting the highest correlation (ρ = 0.647, p = 4.7×10⁻⁴, Figure 2A) and visual trajectory mirroring. Subjects B and D were both moderately-strongly correlated (ρ = 0.554, p = 4.1×10⁻s and ρ = 0.527, p = 6.8×10⁻s, respectively; Figure 2B, D) and had more variability in the respective trajectory profiles compared to Subject A. msTBI Subject C (Figure 2C) had the lowest coefficient value, though still moderately correlated (ρ = 0.447, p = 0.025), likely due to a less mirrored temporal profile between plasma and capillary blood.

**Figure 2.**
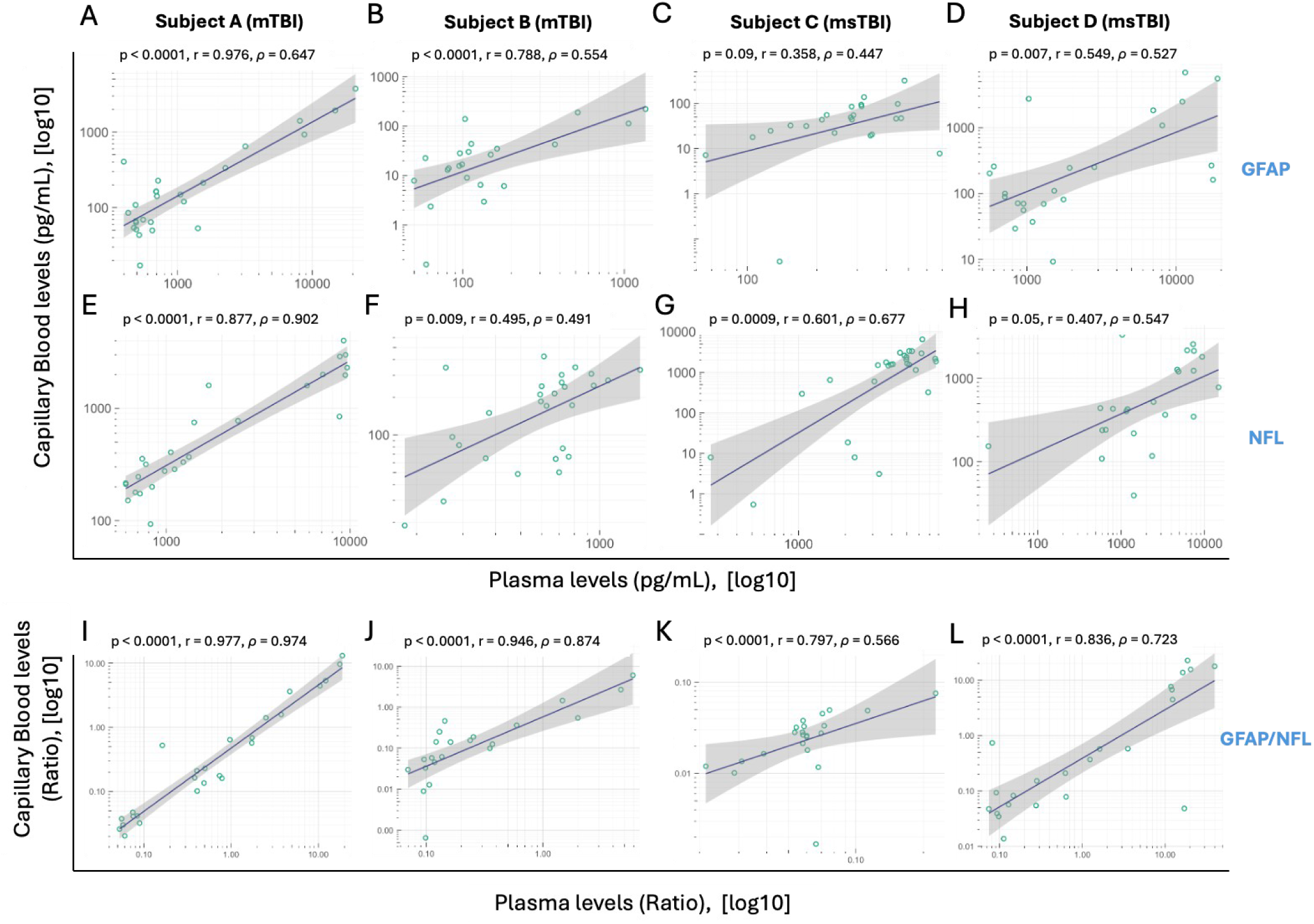
Correlation analysis of biomarker levels for paired venous plasma and capillary blood samples from mTBI and msTBI patients from Prospective Cohort #1. Each point represents results from paired venous plasma and capillary blood samples for GFAP (top row) and NfL (bottom row) levels with daily sampling for the first 48 hours and twice daily sampling thereafter until 14 dpi. (**A, E**) A strong correlation between venous plasma and capillary blood biomarker levels Spearman rho correlation coefficients of r = 0.647 for GFAP and r = 0.902 for NfL was observed for mTBI Subject A. (**B-D, F-G**) Moderate to strong correlations ranging from r = 0.407 to r = 0.788 were observed for Subjects B, C, and D. (**I, L**) The strongest correlations for all subjects were observed for the GFAP/NfL ratio with r=0.797 to 0.977. P-values show significance (p <0.05) for all correlations, except ( **C**). Spearman rho correlation coefficients and p-values are shown with line of best fit and 95% confidence interval.

Similarly, NfL levels showed moderate to very strong correlations, ranging from ρ = 0.407, p = 0.044 (Subject D, Figure 2H) to ρ = 0.902, p = 7.4×10⁻¹⁰ (Subject A, Figure 2E). The lower correlation observed in Subject C (ρ = 0.447, p = 0.025 for GFAP; ρ = 0.677, p = 2.0×10⁻⁴ for NfL) was primarily driven by biomarker levels rising more rapidly in capillary blood than in venous plasma, coupled with missing baseline capillary measurements during the first 48 hours post-injury. Correlation strength was not associated with TBI severity in this cohort.

Importantly, the GFAP/NfL ratio showed higher paired-sample rank correlations for all subjects, with ρ values ranging from 0.797 (p < 10⁻⁵) t) to 0.977 (p < 10⁻¹⁰)) (Figure 2I-L), consistent with our observation in Figure 1. The p-values are either stronger or the same for the GFAP/NfL ratio than for GFAP or NfL alone. This pattern is consistent with the ratio acting as an internal normalization strategy in which a shared per-sample recovery factor cancels in the numerator and denominator; however, ratios of two noisy measurements can also introduce spurious correlation, and we therefore treat the ratio-based improvement as descriptive rather than as evidence of validated bias correction. Formal validation with Bland–Altman limits of agreement and Lin’s concordance correlation coefficient (CCC) will be required before the ratio can be adopted as an assay-quality metric.

### Cross-Sectional Biomarker Profiling (Cohort #2)

Next, to assess diagnostic and monitoring use cases (analytical feasibility) across distinct clinical phenotypes and temporal stages, in this pilot study, we evaluated single timepoint matched samples across varying TBI severities and chronicity (total of 29 TBI subjects, and 21 controls), with 14 geriatric acute civilian TBI subjects, 10 geriatric civilian/Veteran subacute/chronic TBI subjects and 5 Veteran chronic TBI subjects, from three distinct enrollment sites. Only n = 7 GFAP and n = 10 NfL controls are shown because biomarker levels were too low to detect with the DPS device (Table 2). We intentionally focused on older population (mean age 76.1, 73.7 and 47.8 yr old for the three subcohort, respectively), as we envision that the use of the capillary DPS device for biosampling will particularly benefit older populations by reducing the burden associated with sample collection and improving access to biomarker testing in the future.

Paired-sample counts for the Cohort #2 cross-sectional comparison were n = 29 for GFAP and n = 30 for NfL in both TBI and control subjects. Of these, n = 25 subjects had complete paired capillary-venous measurements for both analytes simultaneously in TBI samples (the remainder had single-analyte completeness due to volume constraints), and the Cohort #2 cross-sectional correlations reported below were computed on this n = 25 fully-paired subset for like-with-like comparison. Because serial within-subject samples in Cohort #1 (n = 4) are temporally autocorrelated, the per-subject p-values below should be read as exploratory descriptors of within-subject association rather than as conventional inferential tests (see Limitations). GFAP measurements distinctly clustered according to injury (Figure 3A): acute geriatric samples exhibited the highest GFAP levels (peaking within 24 hours) in the upper right quadrant, whereas chronic Veteran and control subjects clustered in the lower segments, and subacute/chronic geriatric civilian and Veteran patients clustered in the middle of the plot. Paired GFAP samples in both capillary DPS and venous plasma demonstrated moderate linear correlation (Pearson R² = 0.842, p = 1.1×10⁻¹⁰; Spearman r = 0.431, p = 0.031; n = 25 paired samples).

**Figure 3.**
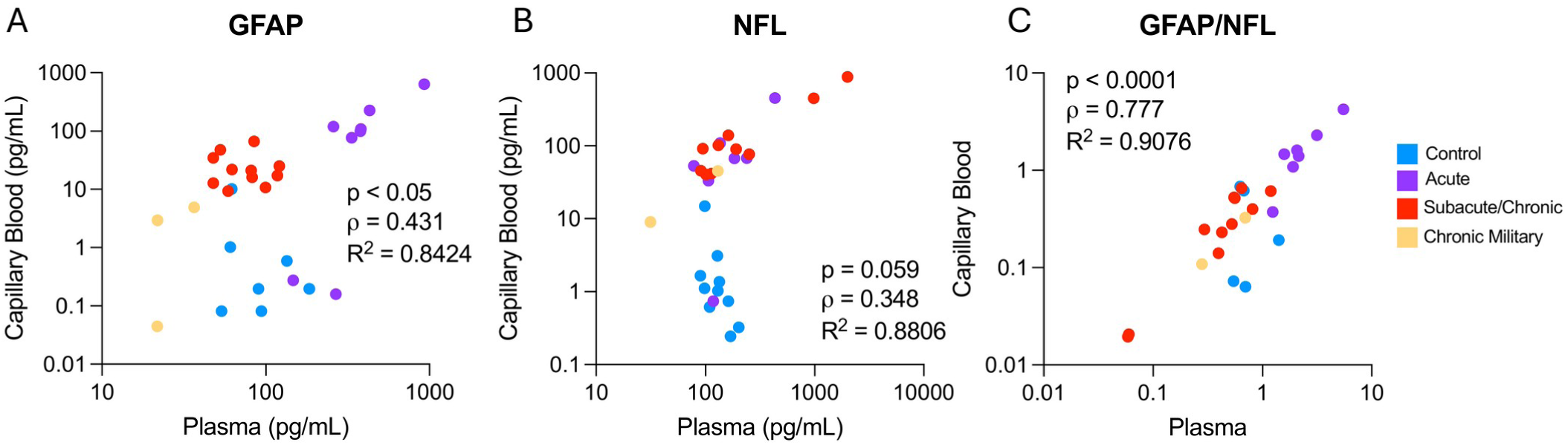
Correlation analysis of biomarker levels from venous plasma and capillary blood collected on a DPS device from a subset of samples from Prospective Cohort #2. Each point represents a paired venous plasma and capillary blood sample, recovered from a DPS device, for GFAP and NfL levels from patients of various TBI severities, indicated in the legend. Pearson correlation coefficient show a significant correlation in (A) GFAP (R ^2^ = 0.8424) and (B) NfL (R^2^ = 0.8806). (C) The ratio of GFAP/NfL normalizes the data and improves the correlation coefficient in both Pearson (R^2^ = 0.9076) and Spearman (r = 0.777) correlations.

Similarly, NfL measurements aligned with canonical temporal expectations (Figure 3B), with control and chronic military samples in the lower quadrants and the highest NfL biomarker measurements from the subacute/chronic geriatric civilian and military sub-cohort in the upper right quadrant. Paired NfL samples also showed strong overall scaling agreement (Pearson R² = 0.881, p = 4.1×10⁻¹²; Spearman ρ = 0.348, p = 0.088; n = 25), indicating that capillary-venous agreement reflects dynamic range across injury phases rather than fine rank ordering.

### Biomarker Normalization using the GFAP/NfL Ratio

Because GFAP and NfL exhibit distinct temporal release profiles post-injury, we evaluated whether the GFAP/NfL ratio behaved as an exploratory internal-normalization metric between capillary and venous samples (n = 25 paired samples; Figure 3C). The ratio showed higher paired capillary–venous correlation than either analyte alone (Pearson r = 0.95, p = 1.6×10⁻¹s; Spearman ρ = 0.78, p = 4.3×10⁻⁶; n = 25). Because GFAP peaks acutely while NfL rises later, the highest ratio values corresponded to acute geriatric samples, as expected from the release biology of the two proteins. This interpretation is also consistent with the well-documented age dependence of these analytes: in TRACK-TBI analyses, plasma GFAP concentrations rise with increasing age and the discriminable dynamic range between injured and non-injured older adults narrows accordingly.^59,60^ Blood-based neuro-biomarkers are likewise altered in older, TBI-exposed populations.^61^ Because our Cohort #2 was intentionally geriatric (mean ages 76.1, 73.7, and 47.8 years across sub-cohorts), age-related baseline elevation of GFAP is expected to contribute to the acute-geriatric ratio maxima observed here; we did not formally test the ratio as a diagnostic classifier in this pilot. Consistent with our findings in Cohort #1, the GFAP/NfL DPS ratio in Cohort #2 was also more strongly correlated between capillary and venous compartments than either analyte alone, reinforcing the value of ratio-based internal normalization across both longitudinal and cross-sectional sampling designs.

## Discussion

### Validation of Dried Capillary Blood Sampling for Neurotrauma Biomarkers

The findings presented here indicate that minimally invasive fingerprick collection of capillary blood — processed with paper-based microfluidic dried plasma spot (DPS) devices — is analytically feasible for GFAP and NfL measurement in TBI and warrants formal validation. Paired capillary DPS and venous plasma measurements showed correlated levels for both astrocytic (GFAP) and neuronal (NfL) markers, though absolute capillary concentrations were lower and more variable than venous plasma. While absolute protein recovery from capillary DPS devices was lower and more variable than traditional venipuncture, temporal trajectories and paired measurements of GFAP and NfL demonstrated moderate to strong correlations (GFAP: Spearman’s ρ = 0.447– 0.647; NfL: ρ = 0.407–0.902). Capillary-based sampling is sensitive to pre-analytical context (including recent exertion), reinforcing the need for standardized collection instructions in remote settings.^36^

The GFAP/NfL ratio showed higher paired capillary–venous correlation than either analyte alone in Cohort #2 (Pearson r = 0.95, p = 1.6×10⁻¹s; Spearman ρ = 0.78, p = 4.3×10⁻⁶; n = 25 paired samples). This pattern is consistent with the ratio acting as an internal-normalization metric in which a shared per-sample recovery factor cancels in numerator and denominator; however, ratios of two noisy measurements can also generate spurious correlation, and we therefore describe the ratio-based improvement as a candidate normalization approach that requires formal validation with Bland–Altman limits of agreement and Lin’s concordance correlation coefficient before adoption.

### Temporal Trajectories in High-Frequency Acute Monitoring

To our knowledge, Prospective Cohort #1 represents the first investigation utilizing direct-to-DPS capillary blood sampling for high-frequency, twice-daily monitoring over a 14-dpi period. This serial sampling accurately captured canonical biomarker trajectories. As expected, GFAP levels peaked acutely within the first few days before sharply declining and plateauing around 6 dpi. Conversely, NfL levels exhibited a gradual, sustained rise throughout the 14-day window. Interestingly, one msTBI patient (Subject C) displayed a modest secondary rise in GFAP after the initial 6-dpi decline. Possible explanations include secondary injury, sampling variability, or previously described prolonged elevation of GFAP or GFAP breakdown products in severe injury32; this single-subject observation is hypothesis-generating only.

### Stratification Across Clinical Phenotypes

In Prospective Cohort #2, capillary DPS GFAP and NfL measurements grouped in a pattern consistent with published release biology across acute geriatric, subacute, and chronic TBI populations (Cohort #2, n = 29). The GFAP/NfL ratio showed the largest between-group separation in this cross-sectional dataset, consistent with the distinct temporal profiles of the two proteins; formal diagnostic stratification was not tested here and would require a prespecified classifier evaluation with outcome linkage.

### Contextualizing our DPS biomarker study Against Current Literature

Several groups have laid the groundwork for microsampling-based neuro-biomarker testing (Figure 4 and Table 3), but important gaps remain. Dried blood spots can enable remote logistics, though hematocrit effects and cellular matrix interference complicate quantification for some proteins.^38,39^ Plasma-separation cards (DPS) reduce these issues by isolating a plasma-like fraction, although many validation studies have relied on venous blood pipetted onto cards rather than true fingerstick collection.^35,37^ More recently, Coleman and colleagues validated remotely shipped finger-prick microtainer samples for NfL (and exploratory GFAP/tau) across multiple neurologic diseases, highlighting both the promise of home sampling and the vulnerability of some analytes to delayed processing (Table 3).^49^ In parallel, the DROP-AD consortium extended capillary DPS/DBS into a multicenter AD setting—including an unsupervised self-collection arm—and reported strong concordance for p- tau217 with supportive agreement for GFAP and NfL between capillary cards and venous plasma (Table 3).^48^ In neurotrauma, Kolanko et al. showed moderate single-timepoint agreement between capillary and venous GFAP and NfL in a cohort that included acute TBI and dementia.^33^ Building on these foundations, our work adds high-frequency, paired trajectories and suggests that the novel approach of utilizing the GFAP/NfL ratio can help normalize collection-related variability. Although our ∼12-hour cadence may miss the exact GFAP apex in some cases, it captured the broader recovery curve; shorter intervals (e.g., every 6 hours) could further refine acute peak timing in future critical-care studies.

**Figure 4.**
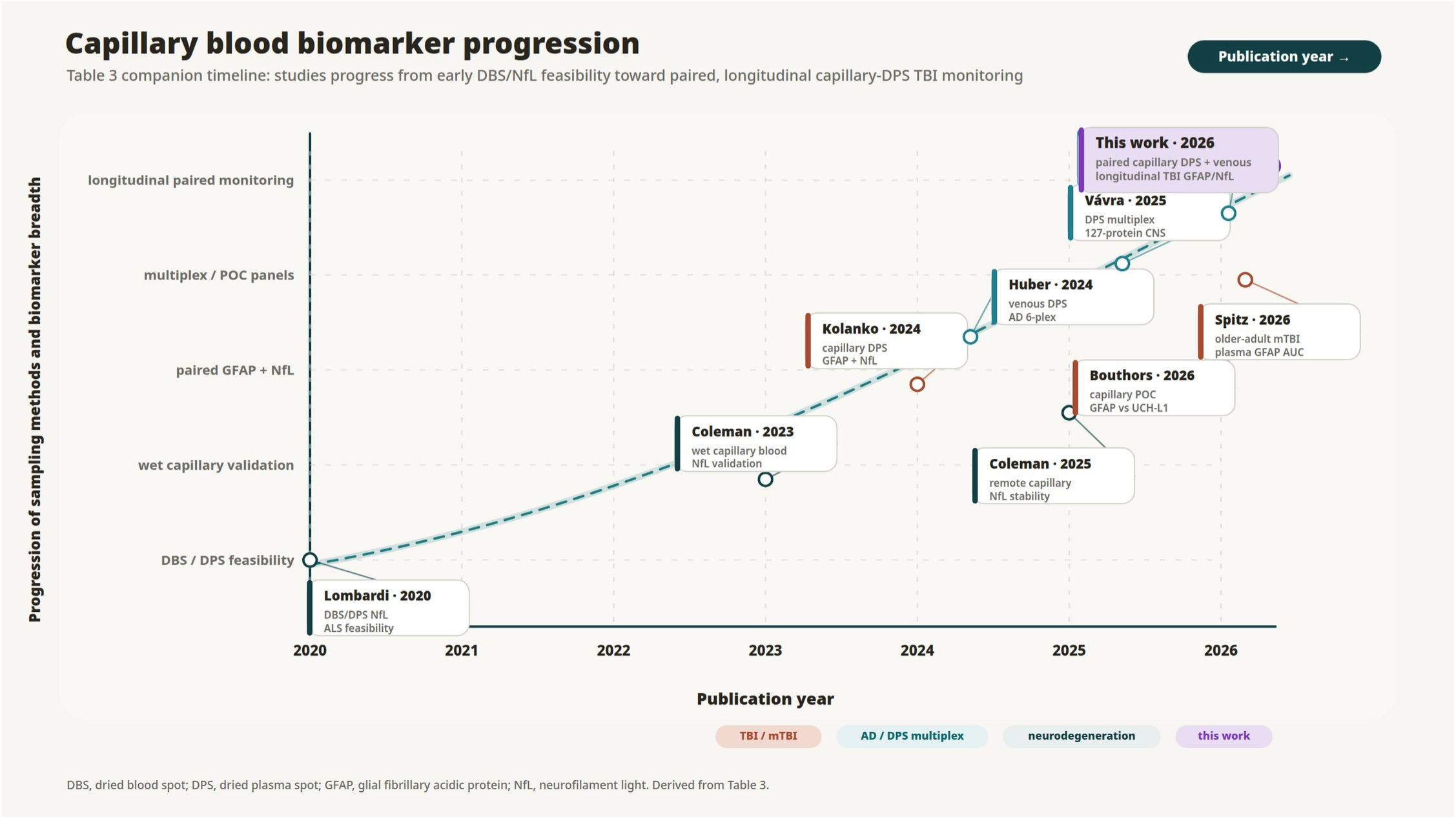
Timeline of selected capillary dried blood / plasma spot biomarker studies in brain health, 2020-2026. Studies are color-coded by primary disease focus (TBI/mTBI in terracotta; Alzheimer’s / DPS multiplex in teal; general neurodegeneration in dark teal). This work (purple, top right) provides the first high-frequency longitudinal (twice-daily x 14 days) paired capillary-DPS and venous-plasma GFAP and NfL trajectories in acute TBI, with cross-sectional validation across acute, subacute, and chronic TBI populations (Cohort #2, n = 29). Companion to Table 3.

**Figure 5.**
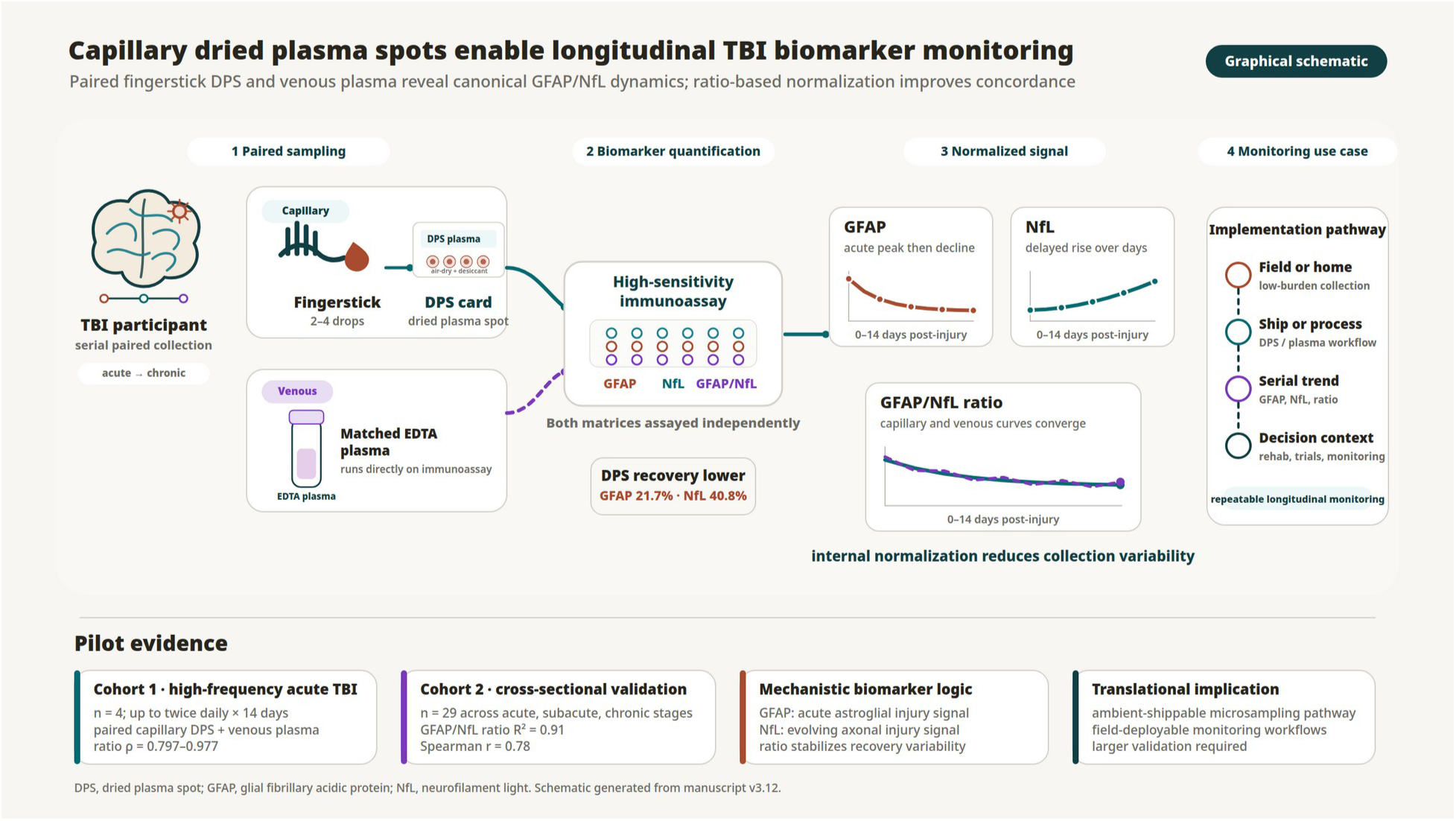
Graphical schematic of the paired capillary dried plasma spot (DPS) and venous EDTA plasma biomarker workflow in traumatic brain injury. Capillary fingerstick blood is processed as DPS, matched venous EDTA plasma is assayed directly, and both matrices are quantified by high-sensitivity immunoassay for GFAP, NfL, and the GFAP/NfL ratio to illustrate longitudinal monitoring from acute to chronic stages.

**Table 3.** Timeline of selected work related to capillary blood studies in TBI or related neurodegenerative conditions (AD, MCI, ALS, HD, PD). *Abbreviations: Cap DPS = capillary dried plasma spot; Ven DPS = venous dried plasma spot; Spearman corr. = Spearman rank correlation coefficient (ρ); AUC = area under the receiver operating characteristic curve; AAUC = age-adjusted AUC; POC = point-of-care*

| Authors | Title | Year | Notes | Subjects (n) |
| --- | --- | --- | --- | --- |
| **This work** | This work | 2026 | Collection device: Telimmune (Noviplex); Collection device: Capitainer SEP10; Capillary blood collected on DPS devices; Venous plasma; Twice daily sampling for 14 days; Acute, subacute, chronic, military, geriatric samples | n = 2 mTBI and 2 msTBI (Cohort #1); n = 29 TBI (Cohort #2); n = 33 total |
| **Huber, Montoliu-Gaya, Brum, et al.<sup>48</sup>** | A minimally invasive dried blood spot biomarker test for the detection of Alzheimer's disease pathology | 2026 | Capillary DPS/DBS collected by fingerstick using Capitainer SEP-10, Capitainer B50, and Telimmune Plasma Separation Card; measured p-tau217, GFAP, and NfL; 337 participants across 7 European centers (304 paired capillary and venous); p-tau217 Spearman rS = 0.74 vs venous plasma; included a self-collection substudy. | n = 337 (CU, MCI, AD, non-AD; includes Down syndrome cohort) |
| **Coleman et al.<sup>49</sup>** | Evaluating finger-prick blood collection for remote quantification of neurofilament light in neurological diseases | 2025 | Finger-prick capillary blood collected into microtainer tubes and processed into plasma/serum; measured NfL and exploratory GFAP and tTau; compared matched venous and capillary samples; simulated 3- and 7-day processing delays (ambient shipment); NfL remained stable, while GFAP was not stable after a 7-day delay. | Discovery cohort: n = 54 controls, 57 HD, 34 MS, 7 ALS, 11 PD; Confirmatory cohort: n = 57 controls, 64 HD |
| **Kolanko et al.<sup>33</sup>** | Quantification of neurofilament light and glial fibrillary acidic protein in finger-prick blood | 2024 | Collection device: Telimmune (Noviplex); Capillary and venous blood DPS; Small TBI cohort; Cap DPS Spearman corr. GFAP = 0.58, NfL = 0.57; Ven DPS Spearman corr. GFAP = 0.43, NfL = 0.57 | n = 19 HC; n = 4 TBI; n = 46 dementia |
| **Coleman et al.<sup>34</sup>** | Validation of remote collection and quantification of blood Neurofilament light in neurological diseases | 2023 | Collection device: Microtainer collection; Wet capillary blood; Measured GFAP, NfL, tTau | n = 23 per group (Control, pre-HD, HD, MS, ALS, PD) |
| **Huber et al.<sup>35</sup>** | Biomarkers of Alzheimer's disease and neurodegeneration in dried blood spots—A new collection method for remote settings | 2024 | Venous blood DPS (Telimmune Plasma Separation Card); measured GFAP, NfL, Aβ40, Aβ42, p-tau181, and p-tau217; DPS correlated with matched EDTA plasma for most analytes; demonstrated diagnostic separation by CSF amyloid status and supported ambient-temperature storage stability for select analytes. | n = 115 AD validation cohort; n = 154 AD discovery cohort |
| **Lombardi et al.<sup>38</sup>** | The potential of neurofilaments analysis using dry-blood and plasma spots | 2020 | Collection device: Telimmune (Noviplex); Collection device: Whatman DBS; Collection device: Protein Saver Card; Measured NfL | n = 47 ALS |
| Bouthors et al.<sup>50</sup> | Capillary UCH-L1 protein blood measurement is affected by sample hemolysis | 2026 | Paired venous + capillary (fingertip/earlobe) blood; Abbott i-STAT Alinity TBI cartridge measured GFAP and UCH-L1; red blood cell hemolysate spiking + human albumin matrix experiments to test hemolysis sensitivity. | Healthy volunteers + neurocritical care patients (covered the clinically relevant concentration range). GFAP: capillary-venous $r = 0.99$ (95% CI 0.98–1.00, $p < 0.001$); UCH-L1: $r = 0.28$ (95% CI 0–0.57, $p = 0.058$) with wide limits of agreement. Hemolysis markedly overestimated UCH-L1 (UCH-L1 is intra-erythrocytic); GFAP remained stable. |
| **Vávra, Traichel, Benedet, Huber, Blennow, Montolieu-Gaya, Ashton, Zetterberg<sup>57</sup>** | Multiplex Biomarker Detection in Dried Plasma Spots: finding the best biomarker for remote blood collection | 2025 | Telimmune Plasma Separation Card; NULISA 127-protein CNS panel; matched DPS vs EDTA plasma; APOe4 $r=0.996$, IL6 $r=0.995$, FABP3 $r=0.994$, p-tau181 $r=0.89$, p-tau231 $r=0.86$, GFAP $r=0.8$, NPTX2 $r=0.92$, NfL $r=0.95$, t-Tau $r=0.93$ | n = 14 discovery cohort (Clinical Neurochemistry Laboratory, Mölndal, Sweden) |
| **Spitz et al.<sup>58</sup>** | Diagnostic accuracy of plasma biomarkers for mild traumatic brain injury in older adults | 2026 | Plasma GFAP, UCH-L1, brain-derived tau, and NfL in older adults with suspected mTBI; age- and sex-adjusted GFAP AUC = 0.93; brain-derived tau AUC = 0.72; age-adjusted cut-offs: GFAP 94–108 pg/mL at age 60 rising to 194–208 pg/mL at age 84; UCH-L1 sex-stratified | n = 89 older adults (60–84 years) |

A complementary finding by Bouthors et al.⁵⁰ has direct implications for the design of capillary biomarker panels. In paired capillary and venous samples assayed on the FDA-cleared Abbott i-STAT Alinity TBI cartridge, GFAP showed excellent capillary-to-venous concordance (r = 0.99; 95% CI 0.98–1.00; p < 0.001); as reported by Bouthors et al., the observed scaling gap remained within their dynamic range without crossing FDA-cleared decision thresholds in that cohort, whereas UCH-L1 showed poor agreement (r = 0.28; 95% CI 0–0.57; p = 0.058) and wide limits of agreement. Bouthors et al. also reported spiking experiments in which hemolysis caused marked overestimation of UCH-L1 across multiple analytical platforms while GFAP remained stable, consistent with the intra-erythrocytic distribution of UCH-L1. For capillary DPS / DBS biomarker panels intended to support point-of-care or remote monitoring workflows, these observations support prioritizing GFAP and NfL — both of which are less sensitive to the preanalytical variability observed in capillary samples and both of which we measured here and found to be analytically robust under DPS conditions — over UCH-L1. Any capillary-specific decision rule would require formal analytical and clinical validation before regulatory use. This is consistent with our own choice to anchor capillary-DPS analyses on GFAP and NfL rather than UCH-L1.

### Implications for Adjacent Neurodegenerative Conditions

Beyond TBI, these proteins are increasingly used to phenotype and stage neurodegenerative disease. In AD cohorts, plasma GFAP tracks amyloid-associated astrocytosis and often rises early in the disease cascade, while NfL provides a complementary readout of neurodegeneration.^40,41^ Multi-analyte panels that combine GFAP and NfL with phosphorylated tau species are now central to blood-based AD research and trial enrichment. ^40,41^ Recent DPS/DBS studies—first using venous blood and now extending to capillary fingerstick collection— suggest that these biomarkers can be measured in remote, low-burden workflows.^35,48^ By demonstrating paired capillary DPS trajectories for GFAP and NfL in TBI, our approach helps bridge neurotrauma monitoring with adjacent aging and dementia applications, particularly where repeated venipuncture is impractical.

### Limitations of the current study

Several additional limitations warrant mention.

This study primarily focused on the development and real-world feasibility implementation of the DPS device, including paired venous blood collection and interrogation of established biomarkers (GFAP and NfL), as well as the novel application of the GFAP/NfL ratio for normalization of DPS-generated measurements. By design, this was a feasibility study and therefore involved a relatively small sample size (n = 33 total (Cohort #1, n = 4 + Cohort #2, n = 29)) across varying TBI severities and chronicity stages. Accordingly, these findings require validation in larger, independent, prospectively collected multi-site cohorts.

All participants in this pilot were adults; pediatric TBI patients, who differ in baseline biomarker levels, hematocrit, and capillary flow characteristics, were not studied, and the applicability of capillary DPS GFAP and NfL to pediatric critical care remains unknown. Additionally, the age and sex sensitivity of capillary biomarker measurements has also become a central concern. Spitz et al. (JAMA Netw Open 2026) showed that GFAP-based mTBI discrimination in older adults (n = 89, aged 60–84) achieves an age- and sex-adjusted AUC of 0.93, but that optimal cut-offs rise from ∼94–108 pg/mL at age 60 to ∼194–208 pg/mL at age 84—a more than two-fold shift—and that UCH-L1 cut-offs differ between sexes.⁵⁸ Their results directly support our prioritization of demographic-aware interpretation of capillary GFAP and NfL, and reinforce the value of the GFAP/NfL ratio, which partially internalizes age-driven baseline shifts and reduces the burden of platform-specific external reference data for prospective deployment.

Although Prospective Cohort 1 included moderate-to-severe TBI cases, we did not systematically link biomarker trajectories to ICU-relevant metrics such as intracranial pressure burden, neurosurgical interventions, or long-term functional outcomes (e.g., GOSE), which precludes conclusions about prognostic or monitoring utility in the neuro-ICU.

The twice-daily longitudinal sampling in four subjects introduces temporal autocorrelation; our within-subject correlation analyses should therefore be considered exploratory and hypothesis-generating rather than definitive inferential tests.

All capillary DPS collections were performed in supervised clinical settings with subsequent frozen storage and central laboratory analysis. Demonstrating usability, ambient-temperature stability, and analytical robustness in unsupervised home, rural, or austere military environments—including collection by non-specialist personnel— will be essential before capillary DPS can be deployed for true point-of-injury or field applications.

A specific design choice in our panel was the prioritization of GFAP and NfL over UCH-L1 as the routine capillary- DPS analytes. Concurrent work by Bouthors et al. (CCLM 2026)⁵⁰ retrospectively supports this analyte selection: UCH-L1 is intra-erythrocytic and therefore sensitive to capillary-collection-related hemolysis on multiple analytical platforms including the FDA-cleared Abbott i-STAT Alinity. The FDA-cleared GFAP + UCH-L1 venous panel should therefore not be expected to translate verbatim to capillary point-of-care or remote-monitoring use; capillary panels should prioritize GFAP and NfL, with UCH-L1 reintroduced only when hemolysis is rigorously controlled at the pre-analytical step.

Beyond UCH-L1, our findings sit within a rapidly evolving capillary-biomarker landscape. The DROP-AD project (Huber, Montoliu-Gaya, Brum, Vávra, …, Blennow, Zetterberg, Ashton et al., Nat Med 2026) showed that dried capillary plasma p-tau217, GFAP, and NfL correlate strongly with venous plasma (rs = 0.74 for p-tau217; r = 0.83 for NfL; r = 0.77 for GFAP) and can discriminate amyloid-positive from amyloid-negative individuals at AUC = 0.86,⁴⁸ supporting the broader feasibility of capillary neuro-biomarker measurement. Multiplex extensions of this approach (Vávra et al., Alzheimer’s & Dementia 2025), using a 127-protein NULISA CNS panel on DPS cards, achieved r > 0.8 against matched venous plasma for the majority of analytes, including GFAP, NfL, p- tau181/217, and total Tau.⁵⁷ These data are consistent with DPS sampling as a substrate compatible with multiple immunoassay platforms for capillary neuro-biomarker measurement, and motivate further prospective evaluation of the GFAP/NfL ratio as an internal-normalization strategy on emerging multiplex platforms.

Although the findings are promising, the limited cohort size may restrict statistical power and generalizability. Larger, prospective studies are needed to further validate these results, particularly to test defined clinical applications prospectively of GFAP, NfL, and the GFAP/NfL ratio for diagnostic, prognostic, and longitudinal monitoring applications in TBI patients.

## Conclusion

This study addresses a critical post-acute monitoring barrier by demonstrating the analytical feasibility and preliminary clinical relevance of capillary blood sampling via dried plasma spot (DPS) devices for GFAP and NfL in TBI. While capillary recovery yields lower and more variable absolute protein concentrations than venipuncture, the temporal trajectories and proportional levels of GFAP and NfL—particularly when applying the novel GFAP/NfL ratio—are strongly correlated with standard venous plasma across both longitudinal and cross-sectional sampling designs. These findings support capillary DPS as a minimally invasive sampling strategy for serial biomarker monitoring in TBI research, including in patients with moderate-to-severe injury, and motivate further development of ratio-based internal normalization to mitigate pre-analytical variability.

If longitudinal capillary biomarker measurement can be established through prospective validation, it may eventually inform how post-acute TBI monitoring is designed in research and clinical care. High-sensitivity cardiac troponin is a useful analogy: building on foundational cardiac troponin assays,42,43 the advent of high-sensitivity assays and validated rapid (0/1-hour) algorithms transformed myocardial infarction triage by enabling rapid, repeatable testing on a scalable platform.^62^ Any analogous role for capillary neuro-biomarkers in neurotrauma will depend on future work linking these measurements to intracranial pressure burden, neurosurgical intervention, and functional outcomes (e.g., GOSE), and on demonstrating usability outside supervised clinical collection. Ultimately, translation will require larger prospective cohorts with linked imaging and functional outcomes, rigorous stability and cost-effectiveness studies for ambient shipment, and clearly defined contexts of use consistent with the FDA-NIH BEST framework. If capillary DPS approaches are to be developed toward potential future in vitro diagnostic applications across TBI, mild cognitive impairment, and Alzheimer’s disease, this will require prospective analytical and clinical validation and independent regulatory review.^46^

Our findings are directly relevant to the biomarker pillar of the recently proposed CBI-M framework for acute TBI characterisation (Clinical, Biomarker, Imaging, Modifiers).^47^ Whereas CBI-M establishes blood-based biomarkers as a formal axis of TBI classification, its biomarker pillar has so far been operationalized largely through venous sampling in acute, supervised settings. Capillary DPS sampling offers a route to operationalize this pillar longitudinally and remotely: the paired capillary–venous concordance for GFAP and NfL demonstrated here — and the GFAP/NfL ratio as a candidate internal-normalization metric — could allow the biomarker pillar to be revisited serially across the acute-to-chronic continuum rather than captured only at presentation. Notably, the CBI-M Modifier pillar explicitly incorporates factors such as age that influence biomarker interpretation, ^47^ reinforcing our observation that demographic-aware interpretation of capillary GFAP and NfL will be essential. Realizing this role will require prospective validation linking capillary biomarker trajectories to imaging, functional outcomes, and the CBI-M pillars in appropriately powered cohorts.

## Declarations

### Ethical Oversight

Ethical oversight covered the entire study and all archived and prospectively collected cohorts, human samples, and data. De-identified archived samples from historical cohorts were obtained from previous observational studies approved by the Institutional Review Board (IRB). Prospective Cohort #1 was approved at Virginia Commonwealth University (VCU) under Advarra protocol Pro00070062. Prospective Cohort #2 was approved under University of California San Francisco (UCSF) protocol 19-28925, which also served as the IRB of record for the University of Pittsburgh, whose local protocol STUDY20110253 ceded review to UCSF; University of Florida protocol IRB202102196 (North Florida Foundation for Research and Education; University of Florida / Malcolm Randall Veterans Affairs Medical Center site); and VA Puget Sound IRB 2 protocol 1653649-1 (Seattle Institute for Biomedical and Clinical Research; VA Puget Sound site).

### Informed Consent

Deidentified, prospectively collected samples were obtained with written informed consent or waiver of consent for all subjects per IRB-approved protocols.

### Consent for Publication

Not applicable (de-identified data only).

### Data Availability

De-identified data from this study are available upon reasonable request to the corresponding author, subject to institutional data sharing policies and patient privacy protections.

### Clinical Trial Registration

This was an observational feasibility study of paired capillary dried plasma spot and venous plasma biomarker sampling; no intervention was prospectively assigned to participants. Clinical trial registration therefore does not apply.

### Authors’ Contributions

DJ, KT, KKW, AD, and WEH conceptualized the work, DJ and KT developed the methodology, DJ and KT led the analysis of biomarkers in the deidentified samples, DJ and WEH performed the formal data analysis, DJ, KT, KKW, and WEH led manuscript writing with assistance from CP and KR, EP, AP, RG, JBW, ABW, LM, and GM.

## Acknowledgments

The authors thank the patients and families who participated in the original observational studies. We thank the clinical research coordinators and laboratory personnel who collected and processed samples. AI-assistance disclosure: The authors used Perplexity Computer (Perplexity AI, San Francisco, CA) as an AI research assistant for literature synthesis, tracked-change reconciliation across coauthor revisions, figure drafting, and manuscript-preparation support during drafting of this manuscript. All scientific content, biomarker data analyses, interpretations, conclusions, and final wording were reviewed, verified, and approved by the human authors, who take full responsibility for the integrity and accuracy of the work.

